# Total parasite biomass but not peripheral parasitaemia is associated with endothelial and haematological perturbations in *Plasmodium vivax* patients

**DOI:** 10.1101/2021.03.19.21253933

**Authors:** João L Silva-Filho, João CK Dos-Santos, Carla Judice, Dario Beraldi, Kannan Venugopal, Diogenes Lima, Helder Nakaya, Erich EV Paula, Stefanie CP Lopes, Marcus VG Lacerda, Matthias Marti, Fabio TM Costa

**Affiliations:** Laboratory of Tropical Diseases – Prof. Luiz Jacintho da Silva, Department of Genetics, Evolution, Microbiology and Immunology, Institute of Biology, University of Campinas, Campinas, Brazil; Wellcome Centre for Integrative Parasitology, Institute of Infection, Immunity & Inflammation, University of Glasgow, Glasgow, UK; School of Pharmaceutical Sciences, University of São Paulo, São Paulo, Brazil; Department of Clinical Pathology, School of Medical Sciences, University of Campinas, Campinas, Brazil; Tropical Medicine Foundation Dr. Heitor Vieira Dourado, Manaus, Brazil; Institute Leônidas & Maria Deane, Fiocruz, Manaus, Brazil

**Keywords:** *Plasmodium vivax*, malaria parasite, total biomass, tissue infection, endothelial activation, haematopoiesis.

## Abstract

*Plasmodium vivax* is the major cause of human malaria in the Americas. How *P. vivax* infection can lead to poor clinical outcomes, despite low peripheral parasitaemia remains a matter of intense debate. Estimation of total *P. vivax* biomass based on circulating markers indicates existence of a predominant parasite population outside of circulation. In this study we investigate associations between both peripheral and total parasite biomass and host response in vivax malaria. We analysed parasite and host signatures in a cohort of uncomplicated vivax malaria patients from Manaus, Brazil, combining clinical and parasite parameters, multiplexed analysis of host responses and *ex vivo* assays. Patterns of clinical features, parasite burden and host signatures measured in plasma across the patient cohort were highly heterogenous. Further data deconvolution revealed two patient clusters, here termed Vivax^low^ and Vivax^high^. These patient subgroups were defined based on differences in total parasite biomass but not peripheral parasitaemia. Overall Vivax^low^ patients clustered with healthy donors and Vivax^high^ patients showed more profound alterations in haematological parameters, endothelial cell (EC) activation and glycocalyx breakdown and levels of cytokines regulating different haematopoiesis pathways compared to Vivax^low^. Vivax^high^ patients presented more severe thrombocytopenia and lymphopenia, along with enrichment of neutrophils in the peripheral blood and increased neutrophil-to-lymphocyte ratio (NLCR). When patients’ signatures were combined, high association of total parasite biomass with a subset of markers of EC activation, thrombocytopenia and lymphopenia severity was observed. Finally, machine learning models defined a combination of host parameters measured in the circulation that could predict the extent of parasite infection outside of circulation. Altogether, our data show that total parasite biomass is a better predictor of perturbations in host homeostasis in *P. vivax* patients than peripheral parasitaemia. This supports the emerging paradigm of a *P. vivax* tissue reservoir, in particular in the hematopoietic niche of bone marrow and spleen.

## Background

Malaria remains a heavy burden across endemic regions worldwide. In 2018 *P. vivax* infection accounted for 41% of all malaria cases outside of Sub-Saharan Africa, with a total of 6.5 million cases and more than 2 billion people in 90 countries at risk ^1^. There are concerns that *P. vivax* elimination will be significantly more difficult than *P. falciparum*, as the current measures for malaria control are less effective for *P. vivax* than for *P. falciparum*, with the elimination of the former presenting a major challenge in areas that successfully reduced *P. falciparum* burden. This persistence is due to some unique biological features complicating treatment and elimination, including low peripheral parasitaemia and presence of dormant liver stages (hypnozoites) which relapse weeks or months after blood infection has been cleared.

*P. vivax* infection is associated with low peripheral parasitaemia (< 2%), as a result of a strict host cell tropism to immature reticulocytes ^2, 3^ that are exceedingly rare in peripheral blood (< 2%) but highly prevalent in the hematopoietic niche of BM and spleen ^4, 5^. Because of limited microvascular adherence *in vivo* and endothelial cell (EC) binding *in vitro* ^6, 7^, it was generally assumed that peripheral parasitaemia reflects the majority of *P. vivax* parasites during infection. However, discrepancy of parasite biomass based on systemic biomarkers such as *Plasmodium* lactate dehydrogenase (pLDH) compared to peripheral parasitaemia supports existence of a major *P. vivax* reservoir outside of circulation, in particular in patients with complicated outcomes ^8, 9^. In support of this hypothesis, studies have demonstrated that late asexual blood stage *P. vivax* parasites (i.e., schizonts) are capable of cytoadhering to endothelial host receptors ^10, 11^ and present at reduced abundance compared to the other blood stages in the blood of *P. vivax* patients ^12, 13^. In experimentally infected non-human primates (NHPs), a significant enrichment of sexual stages (gametocytes) and schizonts in bone marrow sinusoids and parenchyma has been observed ^12^, supporting previous evidence from multiple case reports that identified *P. vivax* in bone marrow and spleen ^14–21^. *P. vivax* parasites can elicit a potent host response, including inflammation and endothelial cell (EC) activation, and cause severe and fatal manifestations at significantly lower peripheral parasitaemia than the more virulent species, *P. falciparum* ^8, 22^. However, the pathogenic mechanisms underlying these alterations in host homeostasis and their relationship with *P. vivax* biomass are not fully understood ^23–26^.

Here we systematically investigated host responses in a cross-sectional cohort of uncomplicated *P. vivax* patients from Manaus, in the Brazilian Amazon region. Our analysis revealed an association between alterations in host homeostasis, including EC activation, damage and haematological disturbances, such as thrombocytopenia, lymphopenia and increased neutrophils turnover, with total parasite biomass but not peripheral parasitaemia. These findings are in line with the emerging paradigm of a clinically relevant parasite reservoir outside of circulation and merit systematic investigations of this reservoir in vivax malaria.

## Methods

### Patients

Peripheral blood and plasma samples was collected from 79 patients infected with *P. vivax*, as diagnosed by light microscopy, seen at FMT-HVD and 34 healthy donors (controls). All individuals within the study were from a local vivax malaria epidemic area in the Amazon region of Brazil. All patients included were outpatients that did not meet World Health Organization (WHO) criteria for severe malaria. Diagnosis was further confirmed by quantitative PCR (qPCR) for both *P. vivax* and *P. falciparum*, using previously published nucleotide sequences ^27^. Exclusion criteria were i) under 18 years of age, ii) pregnancy, iii) use of antimalarials, iv) chronic disease, v) medication known to interfere with platelet count/function and vi) smoking.

Anaemia is defined as haemoglobin < 12.5g/dL; haematocrit < 37%; RBCs counts < 4.45x10^6^/*μ*L. Thrombocytopenia is defined as a decrease in platelet counts to below <150,000/*μ*L. Based on platelet levels, patients were grouped into (i) non-thrombocytopenia (NT: platelet counts >150,000/*μ*L), (ii) mild thrombocytopenia (MT: platelet counts 100,000-150,000/*μ*L), (iii) moderate thrombocytopenia (MDT: platelet counts 50,000-100,000/*μ*L), and (iv) severe thrombocytopenia (ST: platelet counts <50,000/*μ*L). Lymphopenia was defined as a lymphocyte count of less than 1,000 cells/*μ*L. Neutropenia was defined as a neutrophil count of less than 1,500 cells/*μ*L and neutrophilia as a neutrophil count of more than 7,000 cells/*μ*L^28,29^.

### Preparation of poor platelet plasma

After signing the informed consent, 20 mL of venous blood were drawn by venepuncture in a syringe with 15% acid citrate dextrose as anticoagulant to minimize *in vitro* platelet activation. Complete blood counts were done within 15 minutes of blood sampling with a Sysmex KX21N counter. Whole blood was centrifuged at 180 g for 18 minutes at room temperature, without brake for gradient formation, to obtain the platelet rich plasma (PRP). PRP was centrifuged at 100 g for 10 minutes for removal of residual leukocytes, and subsequently centrifuged at 800 g for 20 minutes to obtain the platelet pellet. Prostaglandin E1 at 300 nM was used to minimize platelet aggregation. The supernatant was centrifuged at 1000 g for 10 minutes to obtain platelet poor plasma (PPP).

### Multiplex bead array assay

The biomarkers were analysed in thawed plasma with a customized multiplex suspension detection system (R&D Systems) for quantification of the following biomarkers:

a. pro-inflammatory and myelopoiesis-inducing cytokines: TNF-α, IL-1α, IL-1β, IL-6,IL-8, G-CSF
b. endothelial cell (EC) activation and coagulation markers: ICAM-1, VCAM-1, E-selectin, P-selectin, Angiopoietin-1 (Ang-1), Angiopoietin-2 (Ang-2), von Willebrand Factor (vWF-A2), CD40L, PAI-1, ADAMTS13
c. glycocalyx breakdown and EC damage marker: Syndecan-1
d. platelet activation markers: CXCL4 and CXCL7
e. megakaryocyte differentiation-inducing cytokines: thrombopoietin (TPO) and IL-11; and other proteins such as IL-10, L-selectin and SCF.

A representative set of 31 *P. vivax* patients were selected for the multiplex assay (**Figure S1**). These patients were selected to encompass the wide range of peripheral parasitemia present in the cohort (260 to 25,150 infected RBCs/μL) and to match age, gender and other haematological parameters to those that were not selected. Nine healthy donors matched for age and sex were also selected.

### Plasmodium vivax lactate dehydrogenase (PvLDH) ELISA

To measure PvLDH in patient plasma samples, ELISA was performed using a matching pair of capture and detection antibodies (Vista Diagnostics International LLC, Greenbank, WA, USA). Briefly, 96-well microtiter plate was coated with monoclonal anti-pLDH Vivax-specific (clone 3H8) at a concentration of 1μg/mL in PBS (pH 7.4) and incubated overnight at 4°C. The plate was washed and incubated with blocking buffer (reagent diluent) at room temperature for 1h. After washing, samples were added and incubated for 2h. Next, plates were washed and biotinylated anti-PvLDH detection antibody (clone 6c9), diluted in blocking buffer, was incubated for 2h at room temperature, followed by streptavidin-HRP incubation for 20 minutes at room temperature. Plates were washed and incubated for 20 min with substrate solution. Optical density was determined at 450 nm.

### Principal component analysis and K-means hierarchical clustering

Haematological parameters (haemoglobin levels, haematocrit, differential blood cell counts), parasite parameters (peripheral parasitaemia by blood smear, parasite load by qPCR and parasite biomass PvLDH ELISA) and Luminex data (24 biomarkers) from the selected 9 healthy donors and 31 *P. vivax* patients were normalized to avoid variable-specific bias and z-score values were determined. Since the host response is complex and multidimensional (one dimension per Luminex biomarker), we applied dimension reduction and clustering for ease of downstream analysis. For this, all variables were used as input for principal component analysis (PCA) to reduce the dimensionality of data using the *PCA* function in the *FactoMineR* package in R. For visualization of PCA results *ggplot2*, *factoextra* and *corrplot* packages were used. For each principal component (PC), we determined which variables are better represented and the contribution (correlation or loading score) of each variable for each (PC). Investigation of eigenvalues and the percentage of explained variances retained by the PCs demonstrated that the first 10 PCs accounted for the variance of the data (**Figure S2**). However, variables were well represented by the first 2 PCs (Dim 1 and Dim 2), which were therefore retained for further analysis. In parallel, we performed K-means Clustering (k) followed by bootstrapping, which produced the most stable clusters with k = 2 (Cluster 1 = 21 individuals; Cluster 2 = 18 individuals), which seemed to be the most consistent with the data (**Figure 2A, S3A, B and Table S1**). Table S1 also represents cluster stability via bootstrapping. The metrics of interest is jaccard_index which measures the cluster similarity across bootstrap samples. Similar to the above, k=2 gives stable clusters for all configurations (jaccard_index 0.9 and 0.86). Using Monte Carlo Reference-based Consensus Clustering (M3C) analysis (*M3C* function in the *M3C* package in R) indicated that k=2 is the optimal number of clusters when using K-means clustering (Figure S3C), but when determining spectral clusters, different from elliptical k-means clusters, k=3 gives the best number of clusters (**Figure S3E-G**).

**Table 1:** Demographic, parasite and multiplexed microbead-based immunoassay (Luminex) data obtained from the plasma of a representative subset of 31 *P. vivax* patients and 9 healthy donors (controls).

| Parameters | Healthy donors (n=36) | Symptomatic Pv patients (n=79) | P-value<br>(Pv vs control) |
| --- | --- | --- | --- |
|  | Median [IQR 25-75] | Median [IQR 25-75] |  |
| Age | 32 [23-49] | 36 [28-45] | 0.06 |
| Parasitemia (10<sup>3</sup>/mL) | - | 4.29 [1.86-6.62] |  |
| Parasitemia (%) | - | 0.76 [0.57-1.25] |  |
| Parasite load (copies 18s RNA/mL) | - | 26,642 [9,253-522,297] |  |
| PvLDH (O.D.) | - | 0.18 [0.005-0.34] |  |
| Plasma biomarkers | Healthy donors (n=9) | Symptomatic Pv patients (n=31) | P-value<br>(Pv vs control) |
| TNF-α (pg/mL) | 17.2 [11.0-22.3] | 38.4 [30.0-69.6] | <0.0001 |
| IL-1α (pg/mL) | 11.9 [10.0-19.5] | 25.4 [19.8-33.5] | 0.0004 |
| IL-1β (pg/mL) | 12.0 [8.0-12.8] | 21.4 [14.5-27.6] | <0.0001 |
| IL-6 (pg/mL) | 3.0 [2.5-3.7] | 33.4 [7.6-133.1] | <0.0001 |
| IL-8 (pg/mL) | 2.2 [0.6-2.4] | 6.4 [2.7 – 19.9] | 0.0005 |
| IL-10 (pg/mL) | --* | 314 [169-562] | – |
| G-CSF (pg/mL) | 9.485 [9.485-9.485] | 101.5 [33.49-239.6] | <0.0001 |
| L-Selectin (ng/mL) | 326 [287-391] | 481 [386-579] | 0.0019 |
| ICAM-1 (ng/mL) | 323 [260-464] | 634 [456-849] | 0.0026 |
| VCAM-1 (ng/mL) | 819 [623-959] | 2875 [1753-5108] | <0.0001 |
| E-Selectin (ng/mL) | 26.4 [22.5-33.7] | 56.7 [41.5-74.1] | 0.0001 |
| P-selectin (ng/mL) | 17.0 [15.4-20.6] | 22.2 [17.6-25.7] | 0.0621 |
| Angiopoietin-1 (ng/mL) | 0.4 [0.3-0.6] | 0.5 [0.2-0.9] | 0.8874 |
| Angiopoietin-2 (ng/mL) | 1.8 [1.5-2.1] | 4.3 [2.7-5.3] | 0.0003 |
| Ang-2:Ang-1 ratio | 4.2 [2.7-5.6] | 12.14 [2.7-40.2] | 0.03 |
| VWF-A2 (pg/mL) | 126 [120-150] | 218 [199-277] | <0.0001 |
| ADAMTS13 (ng/mL) | 1110 [483-1740] | 776 [572-1328] | 0.5485 |
| PAI-1 (pg/mL) | 78.9 [62.4-96.4] | 112 [69.3-242] | 0.1541 |
| CD40L (ng/mL) | 0.5 [0.4-0.7] | 1.0 [0.7-1.3] | 0.0001 |
| Syndecan-1 (ng/mL) | 1.8 [1.6-2.4] | 3.7 [2.9-6.0] | 0.0003 |
| IL-11 (ng/mL) | 3.5 [2.9-4.3] | 5.7 [4.7-6.4] | <0.0001 |
| TPO (ng/mL) | 2.0 [1.7-2.2] | 3.0 [2.6-3.4] | <0.0001 |
| CXCL4 (ng/mL) | 0.8 [0.6-1.2] | 1.4 [0.7-2.8] | 0.1236 |
| CXCL7 (ng/mL) | 0.4 [0.4-0.5] | 0.73 [0.4-1.7] | 0.1958 |
| SCF (pg/mL) | 47.61 [37.34-89.34] | 45.68 [36.22-61.39] | 0.1594 |

### Correlation plots and heatmap visualization

Heatmaps were created to visualize variable values using R function *Complex Heatmap*. They represent z-scores using row scaling obtained by centring represented variables with the *scale* function, followed by column clustering using average cluster method and Euclidean distance metric in R. The same software was used to determine pairwise Pearson’s correlation coefficients between variables by running the function *cor* in the *ggcorrplot* package and visualized as a correlogram using R function *corrplot* in the *Hmisc* package displaying positive correlations in red and negative correlations in blue using p ≤ 0.01 as a cut-off.

### Recursive partitioning decision-tree classification and machine learning models

We used recursive partitioning decision-tree classification models to evaluate dominant signatures (attributes) predicting a specific outcome. For decision-tree construction we applied the C4.5 algorithm, using the *RWeka*, *caret* (Classification and Regression Training) and *e1071* packages or the *rpart* package in R. First, the library *caret* is used to create a 10-fold training set to train the model. Then, the algorithm implements decision trees (using the J48 method, which is an open-source Java implementation of the C4.5 algorithm) starting with all instances in the same group, then repeatedly divides the data based on attributes until each item is classified. The attribute on which to divide is selected by information gain, a statistical technique for determining which attribute split will most cleanly divide the data. To avoid overfitting, sometimes the tree is pruned back. In parallel, the algorithm performs k-fold cross validation to measure the performance of a given predictive model and indicates which one has the higher accuracy. Here, we used k=10 to yield test error rate estimates that suffer neither from excessively high bias nor from very high variance ^30^. In parallel, features with mean decrease accuracy larger than 6 were used for random forest. In the random forest analysis, a thousand trees were built using R package *randomForest* (version 4.6.14). The normalized additive predicting probability was computed as the final predicting probability. Those selected important features were used for the random forest analysis on the test cohort for model validation.

### Network analysis

The values of each circulating factor measured in the plasma samples, as well as haematological parameters and parasite biomass in healthy donors and *P. vivax* malaria patients were input in the R software (v 3.4.3) to determine pair-wise Pearson’s correlation coefficients to generate correlation networks and the *p*-value to test for non-correlation was evaluated using p ≤ 0.05 as a cut-off. In order to analyse the structure of the networks, edges list was generated in R using the functions *melt* (*reshape2* package), *graph_from_edgelist* (*igraph* package). Graphs were customized in the Cytoscape software (v 3.5.1) using the force-directed layout, which simulates a system of forces, determined by the correlation strength. In the in the equilibrium state edges tend to have uniform length, and nodes that are not connected by an edge tend to be drawn further apart.

### Stimulation of HUVEC with patients’ plasma pools

After standardization procedures, primary human umbilical vein endothelial cells (HUVEC) were stimulated or not (mock control) in culture media for 6h – to evaluate mRNA expression – or for 18h – to evaluate protein expression - with complete EGM-2 medium (Lonza) containing 30% (v/v) plasma pools generated from the three subgroups - Healthy control, Vivax^low^ and Vivax^high^ - and 3U/L heparin.

### Real-Time Quantitative RT-PCR

After 6h stimulation, total RNA was isolated from the cell lysate using the miRVana miRNA Extraction kit (Ambion) according to the instructions of the manufacturer. cDNA was synthesized with TaqMan Reverse Transcriptase (Applied Biosystems, Foster City, CA) and mRNA expression of genes were determined by qRT-PCR. Real-time qRT-PCR was performed on an ABI-Prism 7000 PCR cycler (Applied Biosystems) or on the CFX96 Real-Time PCR Detection System (Bio-Rad). Cycling parameters were 95°C for 1min and then 35 cycles of 95°C (15s) and 60°C (1min), followed by a melting curve analysis. The median cycle threshold (C_t_) value and 2^-ΔΔCt^ method were used for relative quantification analysis, and all C_t_ values were normalized to the GAPDH mRNA expression level. Results expressed as means and SEM of biologic replicates are shown. The mock sample (HUVECs incubated with culture media only) was used as reference. The oligonucleotides used are described in Supplemental Table S3.

### Endothelial cell Flow Cytometry (FC) and Immunofluorescence Analysis (IFA)

For immunofluorescence analysis, cells were grown in 8-well chambered coverslips (IBIDI) until confluence. After 18h stimulation with plasma pools, cells were washed with PBS and fixed/permeabilized with ice cold 100% methanol for 5 minutes at -20°C. In brief, cells were incubated with 10% goat serum (ThermoFisher) to avoid secondary antibody nonspecific binding for 1h at room temperature and then incubated with specific primary antibodies to human ICAM-1 (mouse monoclonal clone MEM-111; Abcam; cat. number ab2213;) (used at a dilution of 1:100 in 10% goat serum); VCAM-1 (mouse monoclonal clone 1.4C3; Abcam; cat. number ab212937) (used at a dilution of 1:500 in 10% goat serum) and mouse IgG1 isotype control (Dako; cat. Number X0931) (used at a dilution of 1:10 in 10% goat serum) overnight at 4°C. After washing, wells were overlaid for 1h with AF488-conjugated secondary antibody (used at a dilution of 1:500 in 10% goat serum) and Hoechst (diluted at 1:2000) at room temperature. For controls, primary antibodies were omitted from the staining procedure and were negative for any reactivity. The chambers were placed at 4°C until use for immunofluorescence assay (IFA).

In flow cytometry, after 18h stimulation with 30% plasma pools, cells were washed 2x with DPBS and treated with Accutase® Cell Detachment Solution (Biolegend) at room temperature for up to 3 minutes, or until the cells are detached. Cell count and viability with trypan blue dye were determined and cells were resuspended in ice cold DPBS without calcium/magnesium, 0.5mM EDTA, 10% foetal bovine serum (FBS) (GIBCO). Cells were incubated with FcBlock (BD Biosciences, San Jose, CA, USA), followed by incubation with unconjugated anti-VCAM (mouse monoclonal clone 1.4C; Abcam; cat. number ab212937) or AF488-conjugated anti-ICAM-1 (mouse monoclonal clone HCD54; Biolegend; cat. number 322714) or unconjugated mouse IgG1 isotype control (Dako; cat. Number X0931) for 1h at 4°C. Cells were then washed and incubated for 1h at 4°C with secondary antibody AF488-conjugated anti-mouse IgG (ThermoFisher). Finally, cells were incubated with Fixable Viability Dye eFluor™ 506 (ThermoFisher) in DPBS without calcium/magnesium, 0.5mM EDTA for 30min at 4°C. Cells were washed and resuspended in buffer and acquired using a BD FACSCelesta cytometer (100,000 events/sample). Percentage of positive cells and expression profiles for ICAM-1and VCAM-1 were then determined by the mean fluorescence intensity using FlowJo software (Ashland, OR, USA).

### Ex vivo Evaluation of Endothelial cell Monolayer Function

Endothelial cell monolayer function was measured using ECIS, an electric cell-substrate impedance sensing system (ECIS Zθ, Applied BioPhysics, Troy, NY), as previously described ^31^. The system then applies weak alternating currents through the electrode array and continuously measures the ability of the cell monolayer to impede the movement of electrons between adjacent endothelial cells (resistance). Briefly, cells were seeded at 2.5 × 10^5^ cells/well on fibronectin-coated (10 µg/ml) eight-well arrays (8WE10, Applied BioPhysics, Troy, NY) containing interdigitated gold electrodes. Endothelial cells were seeded 48 h before experiments and the resistance started to be recorded after 48 h. Only wells with resistance > 1,500 ohms and stable impedance/resistance readings were used. Before stimulation, resistance was continuously monitored for 2 h, to confirm monolayer stability represented by a plateau in the resistance curve. Stimuli (20% v/v pooled plasma in complete medium) was then added to wells under continuous impedance/resistance monitoring for 12h. A baseline resistance value was recorded immediately prior to the addition of each stimuli and results are expressed as a ratio from baseline resistance (normalized resistance).

### Statistical Analysis

Fisher’s exact test was used for categorical data. Data normality was checked by the Shapiro-Wilk test. Student’s t-test was used to compare means between groups with normally distributed data, and data sets with non-normal distributions were compared using the Mann– Whitney test. All tests were performed two-sided, using a nominal significance threshold of *p* < 0.05 unless otherwise specified. When appropriate to adjust for multiple hypothesis testing, Tukey’s or Bonferroni corrected multiple comparisons test significance at the p-value < 0.05 threshold was performed unless otherwise specified. Data are presented as scatter plots with median and 25%-75% interquartile range, box plots showing minimum to maximum range or means and SEM, unless otherwise stated. Analyses were performed and the graphs generated in GraphPad Prism 8 and R software.

### Study approval

All subjects enrolled in the study were adults. Written informed consent was obtained from all participants and the study was conducted according to the Declaration of Helsinki principles. The study was approved by the local Research Ethics Committee at Fundação de Medicina Tropical Dr. Heitor Vieira Dourado (FMT-HVD, Manaus, Brazil), under #CAAE: 54234216.1.0000.0005.

## Results

### Uncomplicated *P. vivax* patients present with haematological changes

We have conducted a cross-sectional study with uncomplicated *P. vivax* malaria patients seen at FMT-HVD in Manaus, Brazil. We included 79 adult patients (median age of 36 years) with confirmed *P. vivax* infection (smear and PCR positive) and 34 age-matched uninfected healthy donors (controls) (**Table 1**). All individuals within the study including controls were from the State of Amazonas, in the Amazon region of Brazil. Blood was collected at enrolment for determination of haematological parameters, peripheral parasitaemia by Giemsa staining of blood smears and PCR to determine genome copy numbers. Preparation of poor platelet plasma (PPP) was done within 15 minutes of sampling. The median peripheral parasitemia was 4,290 infected red blood cells (iRBCs)/μL of blood (25-75 interquartile range 1,860-6,620 parasites/μL) and parasite load of 26,642 copies of 18s RNA/μL (25-75 interquartile range 9,253-522,297). We also measured total parasite biomass independently of peripheral parasitaemia by quantifying levels of *P. vivax* lactate dehydrogenase (PvLDH) in plasma (**Table 1**).

Analysis of haematological parameters revealed significantly reduced haemoglobin levels and haematocrit across *P. vivax* patients compared to controls, with anaemia in 38% of the patients (**Figure 1A**). Similarly, leukocyte numbers were significantly decreased (Mean ± SD: 4.36±1.74 x10^3^/*μ*L vs. 5.72±1.34 x10^3^/*μ*L, *p*=0.0004), with 54.5% of the patients presenting with leukopenia (defined as a leukocyte count < 4000 cells/*μ*L). In contrast, neutrophil counts were not significantly different, and only 8.3% of *P. vivax* patients were presenting with neutropenia (neutrophil counts < 1,500 cells/*μ*L) (**Figure 1B**). Other myeloid cell populations however, such as monocytes, basophils and eosinophils (MXD), were significantly reduced. We also observed a significant reduction in lymphocyte and platelet counts in this cohort (**Figure 1C**), with 80% presenting with lymphopenia (lymphocyte counts < 1,000 cells/*μ*L) and 87% with thrombocytopenia (platelet counts < 150,000 cells/*μ*L), many of them with severely reduced levels (**Figure 1C**). Alterations in platelet counts were accompanied by the release of mega platelets in the peripheral circulation as a significant increase on mean platelet volume was observed (**Figure 1C**).

**Figure 1:**
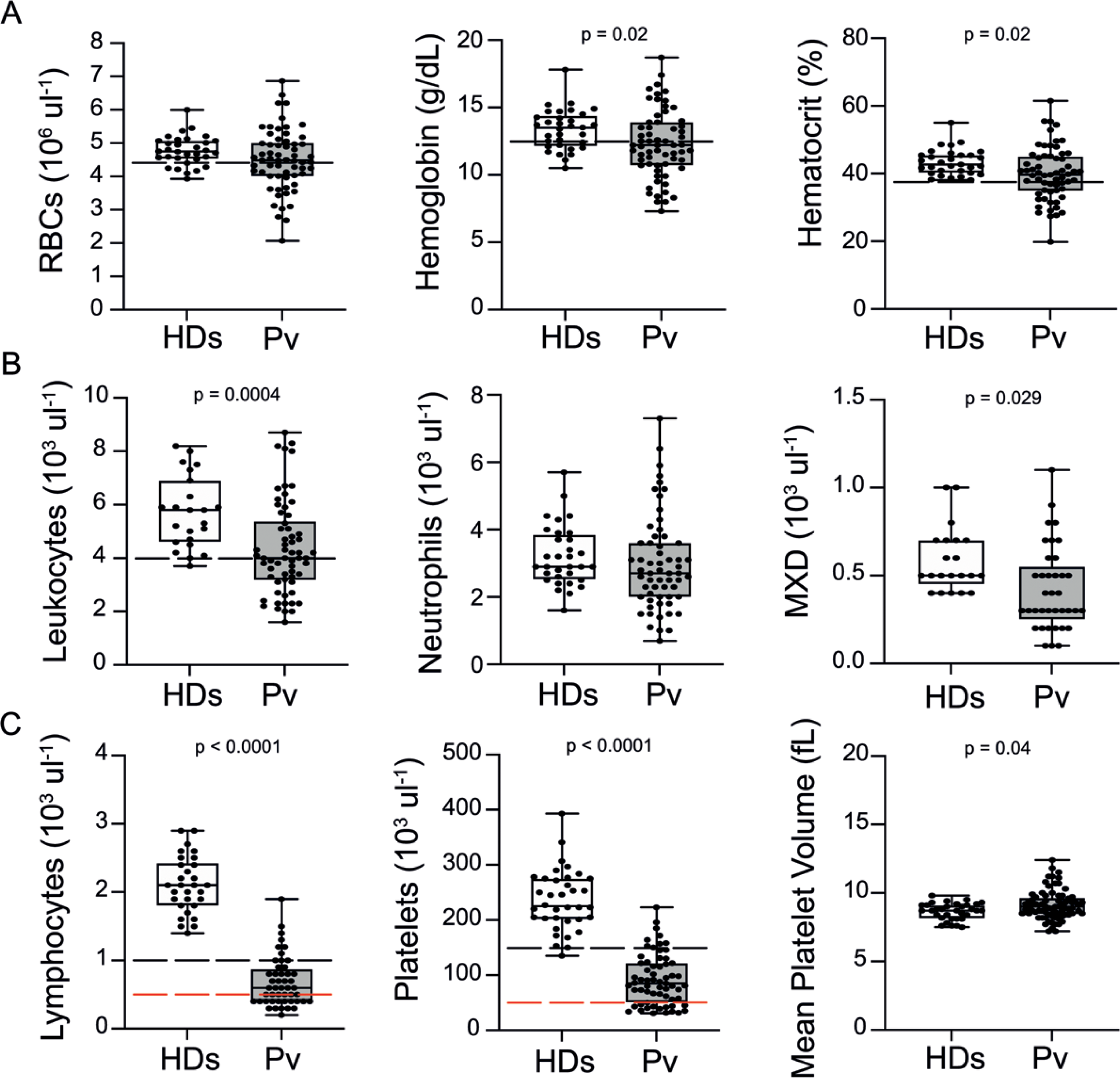
Clinical data of *P. vivax* patients (Pv) and healthy donors (HDs). **(A) Red blood cell parameters.** Shown are red blood cell counts, hemoglobin levels and hematocrit. **(B) Other blood cell parameters.** Shown are numbers of leukocytes, neutrophils, and monocytes, basophils and eosinophils (MXD). **(C) Number of lymphocytes, platelets and mean platelet volume (MPV)**. Parameters are depicted as box plots showing each individual value and the median with maximum and minimum values. Dashed lines in black mark the minimum threshold for normal reference values, while lines in red mark threshold for severe lymphopenia and thrombocytopenia, respectively. Two-tailed student’s t-test was used to compare variables with normally distributed data, and Mann-Whitney test was used to compare variables with non-normal distributions; *p*-value is indicated above the graph when *p* < 0.05. HDs = healthy donors (controls, n= 34); Pv = *P. vivax*-infected patients (n = 79).

**Figure 2:**
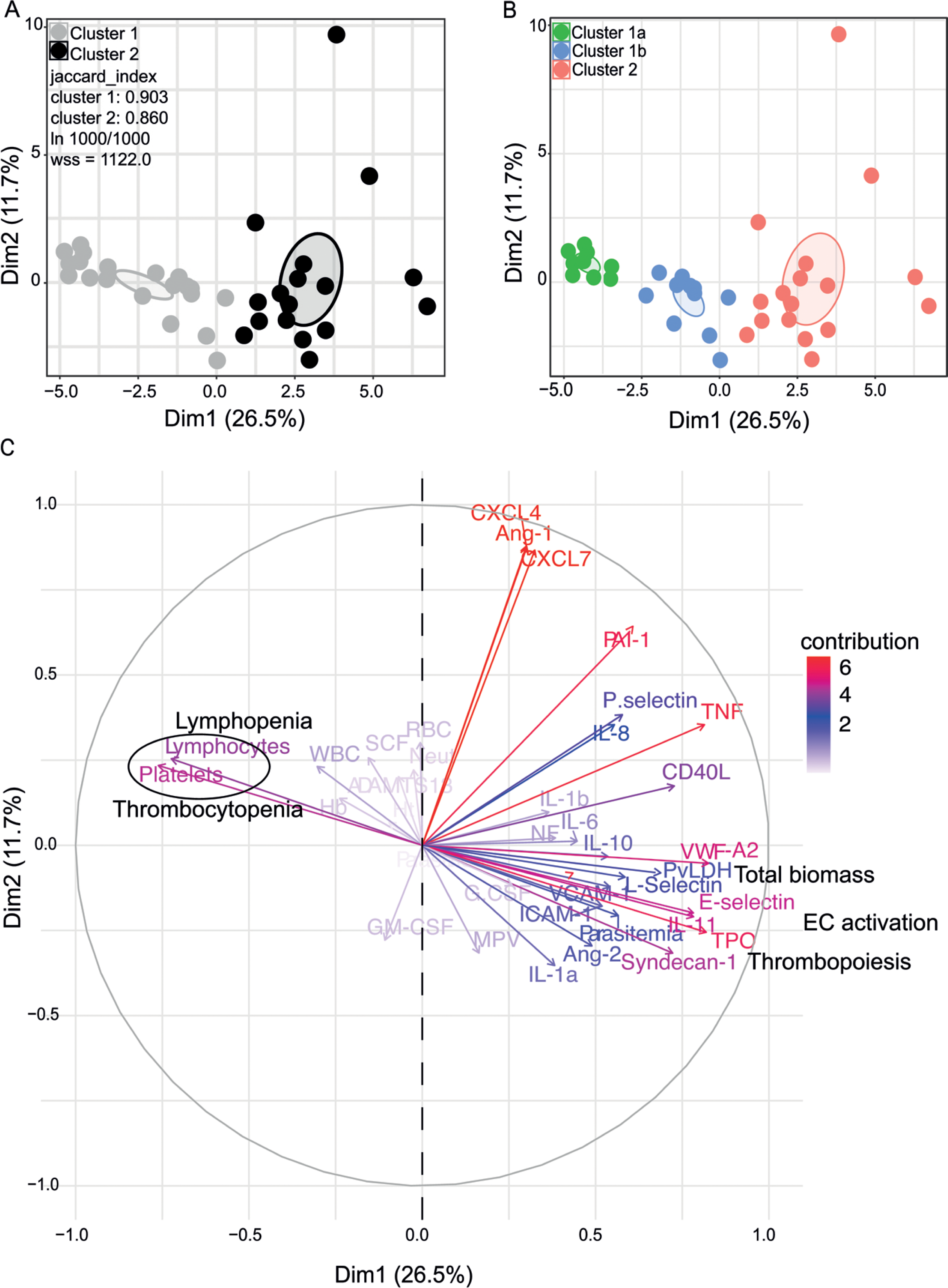
Characterization of heterogeneity in symptomatic *P. vivax* patients defines clusters of individuals. **(A, B) Clustering of patients and healthy controls.** After z-score normalization, Principal Component Analysis (PCA) was performed for data dimensionality reduction. K-means Clustering using k=2 followed by bootstrapping (1,000 times) in a PCA plot was performed and produced the most stable clusters regardless of the starting point (ln 1,000/1,000): Cluster 1 = 23 individuals comprising of 9 healthy donors and 14 *P. vivax* patients and Cluster 2 comprising 17 *P. vivax* patients. The jaccard_index measures cluster similarity across bootstrap samples (jaccard_index ranges from 0 to 1, an index < 0.6 hints at a weak, unreliable cluster while > 0.85 means generally reliable). As indicated in the PCA plot, k=2 gives stable clusters for all configurations (jaccard_index 0.9 and 0.86) and withinss (wss) = 1,122. Open ovals represent 95% confidence interval ellipses around group mean points. Principal Component Analysis (PCA) was performed for data dimensionality reduction, in parallel with K-means Clustering (k) followed by bootstrapping (1,000 times). Open ovals represent 95% confidence interval ellipses around group mean points. (B) The resulting clusters represent healthy controls (1a) and patients (1b, 2). **(C) Contribution of variables to clustering.** In the circular plot the correlation between each input variable and principal components is used as coordinates (loading score). Plots shows how covariables determine patient distribution in the PCA plot.

In summary, patients in our cohort presented with a wide range of parasitaemia and uncomplicated clinical signs of *P. vivax* infection at medical consultation. However, significant haematological abnormalities were present in the majority of patients during early onset of disease, in line with previous findings ^8,23, 28, 32–35^.

### Unsupervised clustering reveals two *P. vivax* patient subgroups that differ in parasite biomass: Vivax^high^ vs Vivax^low^

To determine whether the observed changes were associated with specific host signatures, in particular circulating biomarkers of haematological and endothelial changes, we applied a multiplexed microbead-based immunoassay (Luminex) in a representative subset of 31 *P. vivax* patients and 9 controls, as explained in the Methods section (**Figure S1**). We selected a series of circulating biomarkers associated with haematological changes, including cytokines altering thrombopoiesis (TPO and IL-11), myelopoiesis and lymphopoiesis (TNF-α, IL-1α, IL-1β, IL-6, IL-8, G-CSF) ^36–38^. In addition, we selected markers of endothelial cell (EC) and platelet activation, coagulation (ICAM-1, VCAM-1, E-selectin, P-selectin, Angiopoietin-1 and -2, CD40L, VWF-A2, ADAMTS13, PAI-1, CXCL4, CXCL7) and EC glycocalyx breakdown (Syndecan-1).

We observed significant upregulation of multiple cytokines associated with haematological changes in the *P. vivax* patients compared to control (**Table 1**). In addition, patient samples exhibited a strong phenotype of increased EC activation, glycocalyx breakdown and coagulation. The high interquartile range in parasitaemia and host signatures (**Table 1**) suggested a heterogenous phenotype across the patient population. In order to identify possible stratification of patients into distinct subgroups, we further analysed the clinical data (**Figure 1**), parasite parameters and Luminex data (**Table 1**) from the same 31 *P. vivax* patients and 9 controls as above. After z-score normalization, Principal Component Analysis (PCA) was performed for data dimensionality reduction, considering the large number of variables in our dataset. Next, we ran K-means Clustering (k) followed by bootstrapping (**Figure 2A, B; Figures S2, S3, Table S1**) to identify possible subclusters of individuals. This analysis revealed consistent separation of samples into 2 clusters, one of them including all controls (Cluster 1a) and a subset of 14 patient samples (Cluster 1b) and a second one representing the remaining 17 patient samples (Cluster 2) (**Figure 2A, B**). In order to visualize covariables of the observed patient distribution (PCA) and clustering (K-means), we plotted the correlation (loading score) of each input variable with a principal component (**Figure 2C**, **Table S2**). This analysis demonstrated covariation of lymphopenia and thrombocytopenia on one hand, and markers of EC changes, platelet production, activation and parasite parameters (PvLDH and peripheral parasitemia) on the other hand as major contributors to the principal components (**Figures 2C**). Direct comparison of the 2 patient subgroups revealed significant higher total parasite biomass but not peripheral parasitaemia or parasite load (**Figure 3A**). In agreement with previous findings ^8,9,39^, z-score comparison further demonstrated that total parasite biomass was higher than and not correlated with peripheral parasitaemia levels or parasite load, in particular in patients of cluster 2 (**Figure 3B, C**). In addition, PvLDH was the input parasite variable with the highest loading score (correlation = 0.59) and lowest *p*-value (0.0000917) in the first PC dimension when compared with peripheral parasitaemia and parasite load (**Figure 2C**, **Table S2**).

**Figure 3:**
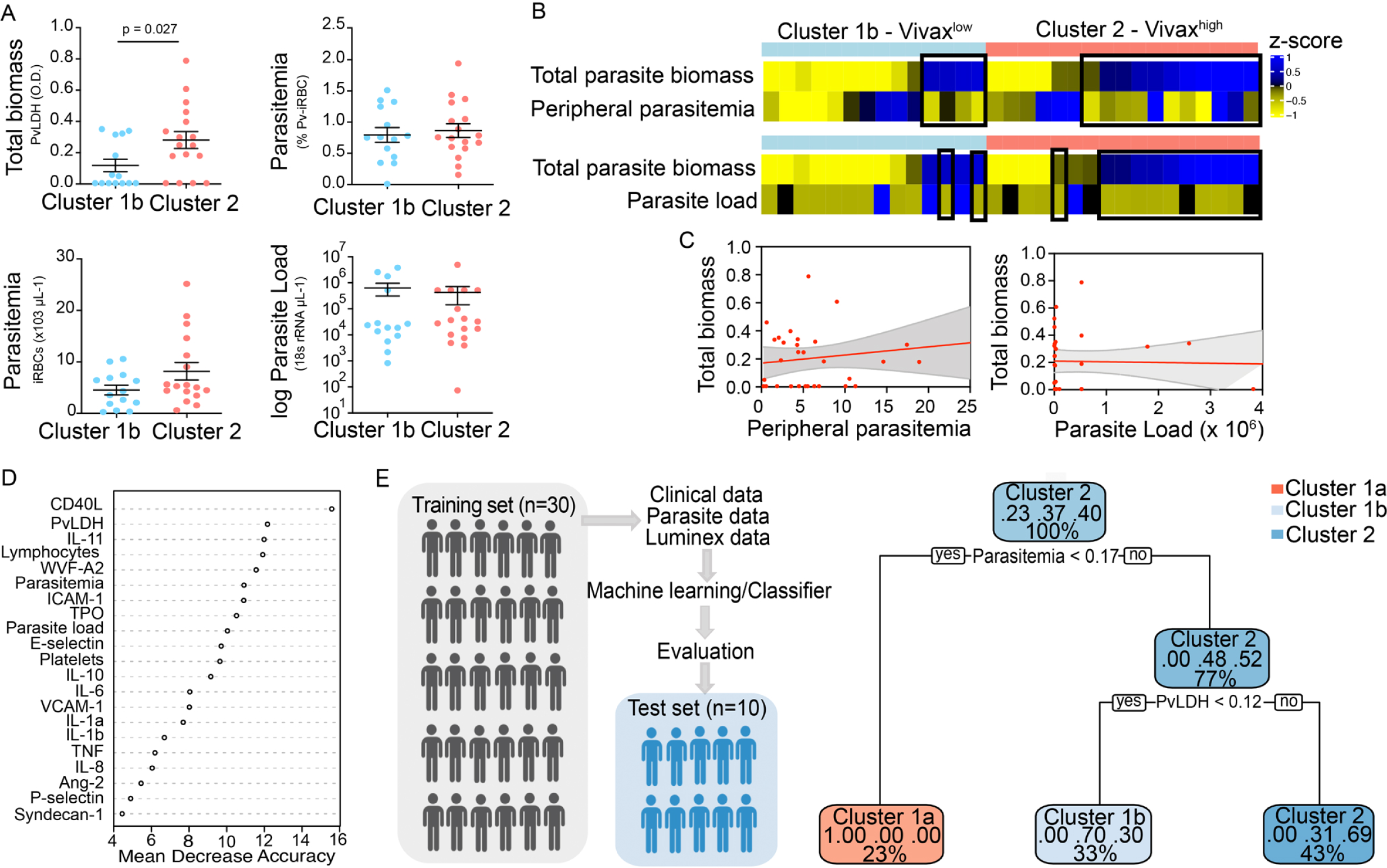
Unsupervised clustering analysis reveals two *P. vivax* patient subgroups that differ in parasite biomass. **(A) Parasite parameters *vs* patient clusters.** Comparison of the two patient clusters (Cluster 1b and Cluster 2) across parasite parameters reveals significant differences with total parasite biomass (PvLDH) but not peripheral parasitaemia or parasite load (copies of 18s rRNA/μL of blood). **(B) Parasite biomass *vs* parasitemia across clusters.** Heatmap represents z-scores of PvLDH with peripheral parasitemia or parasite load, respectively. Black boxes highlight patients with relatively lower peripheral parasitemia compared to PvLDH levels, indicating the underestimation of total parasite biomass based on peripheral parasitemia values. **(C) Correlation between parasite biomass and parasitemia.** Scatter plot showing lack of correlation between PvLDH and peripheral parasitemia or parasite load, respectively. Regression line in red, with 95% confidence interval shown in shaded gray. **(D, E) Predicting parasite clusters. (D**) Top parameters prioritized by random forest analysis ranked by the mean decrease in accuracy. **(E)** Best-fit decision trees and random forest machine learning models corroborate PvLDH value as the most important parasite signature in segregating patients into clusters 1b and 2. Cut-off values of the attribute that best divided groups were placed in the root of the tree according to the parameter value. The total of classified registers for each class and the percentage of observations used at that node are given in each terminal node.

Indeed, using a best-fit classification tree model and a random forest machine learning model defining k-means clusters as categorical outcome, PvLDH is the best parasite predictor attribute segregating patients into clusters 1b and 2 (**Figure 3D, E**). After both models were trained in a set of 30 individuals, randomly selected by the training algorithm set, they were tested in the 10 remaining individuals, where all cluster 1a (control) individuals and 80% of *P. vivax* patients were correctly classified in either cluster 1b or cluster 2. Based on these observations we designated cluster 1a as Control cluster (representing the healthy donors), cluster 1b as Vivax^low^ (representing patients with low *P. vivax* biomass) and cluster 2 as Vivax^high^ (representing patients with high *P. vivax* biomass).

### Different levels of haematological alterations between Vivax^high^ and Vivax^low^ patients

The three clusters were not significantly different in patient age (median: 33; IQ 25-75: 22-57), gender (80% male; 20% female in each cluster), average days of symptoms when samples were collected, haemoglobin levels, haematocrit or RBC counts, indicating that these parameters are not confounders accounting for the differences observed between the clusters (**Figure 4A**). However, systematic analysis of haematological parameters between Vivax^high^ and Vivax^low^ patients revealed significant differences. Vivax^high^ patients showed a more intense reduction in platelet counts when compared to Vivax^low^ patients (Vivax^high^ 63,000 ± 6,413 vs Vivax^low^: 100,700 ± 9,381; p = 0.002), with a higher frequency of patients with severe thrombocytopenia (Vivax^high^ 47% vs Vivax^low^ 8%) (**Figure 4B**). Although not significant, there was a trend in the reduction of lymphocyte counts in Vivax^high^ patients when compared to Vivax^low^, with 88% of Vivax^high^ patients presenting lymphopenia versus 64% in Vivax^low^ patients. In addition, we observed a 4-fold increase in the frequency of patients with severe lymphopenia in the Vivax^high^ cluster compared to Vivax^low^ patients (**Figure 4B**). While there was no change in the number of circulating neutrophils in the different clusters of individuals, mixed cell counts (MXD), a parameter representing monocytes, basophils and eosinophils numbers, was significantly reduced in Vivax^high^ patients. As a result, there was a significant enrichment of neutrophils in the leukocyte fraction in the blood of Vivax^high^ patients as well as a higher Neutrophil:Lymphocyte count ratio (NLCR) (**Figure 4B**).

**Figure 4:**
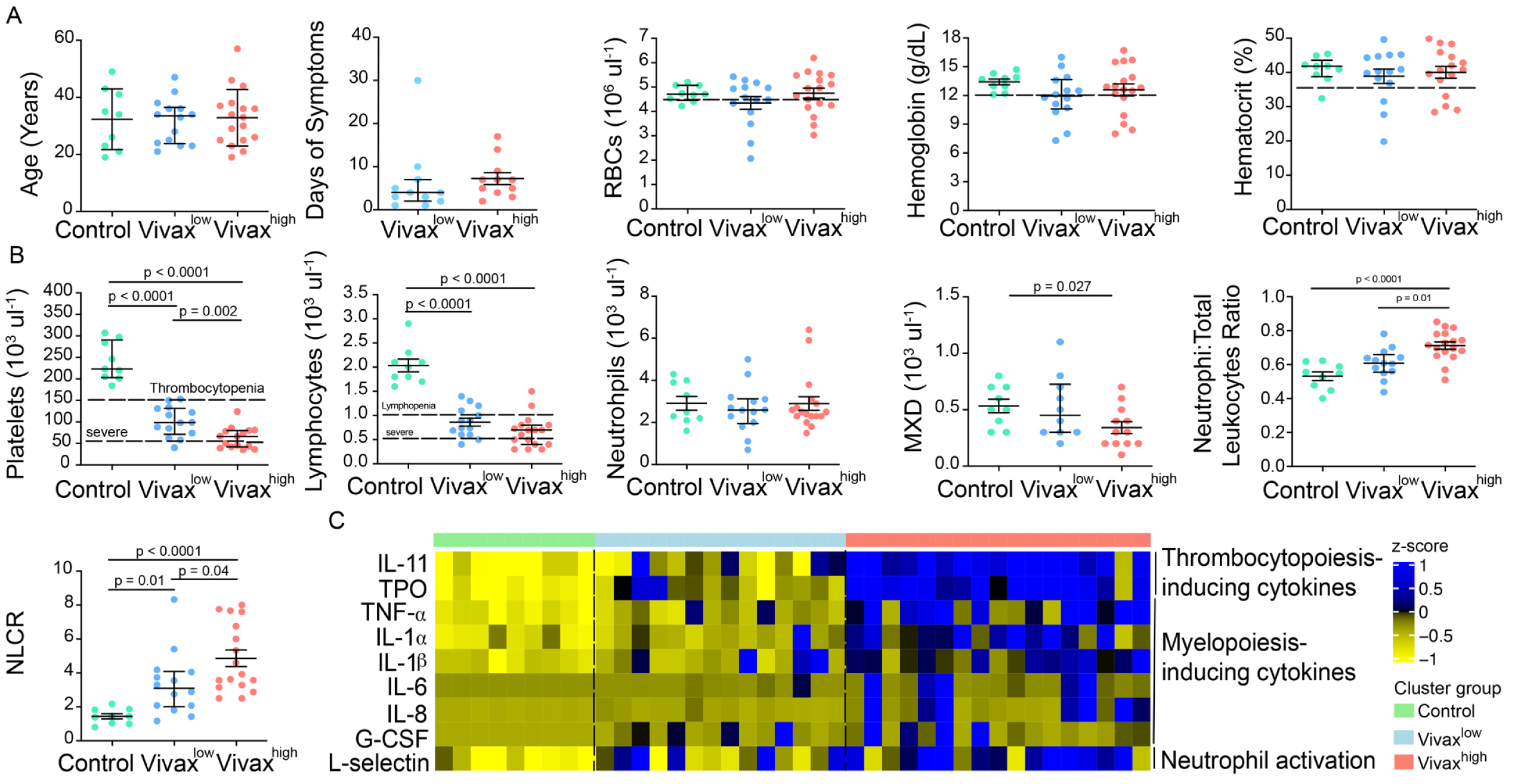
More severe haematological alterations in Vivax^high^ compared to Vivax^low^ patients. **(A) Patient data and hematological parameters.** Comparison of patient age, average days of symptoms when samples were collected, haemoglobin levels, haematocrit or RBC counts across patient clusters (Control: n=9; Vivax^low :^ n=14; Vivax^high^: n=17). Data are depicted as plots showing individual values and the median (black lines) and the interquartile range. **(B) Blood cell counts.** Comparison of differential haematological counts across clusters. Shown are. numbers of platelets, lymphocytes, neutrophils and monocytes, basophils and eosinophils (MXD), neutrophil to total leukocyte ratio and neutrophil to lymphocyte ratio (NLCR). Top dashed lines mark the minimal threshold for normal reference values, while bottom dashed lines mark the threshold for severe lymphopenia and thrombocytopenia, respectively. Parameters are depicted as plots showing individual values and the median (black lines) and the interquartile range. One-way analyses of variance with Bonferroni-corrected multiple comparisons test were performed. *p*-value is indicated above the graph when reached significance of *p* < 0.05. **(C) Cytokine response and neutrophil activation across clusters.** Heatmap represents z-scores obtained by centering values of Luminex data. Shown are thrombopoiesis-inducing cytokines, myelopoiesis-inducing cytokines and neutrophil activation markers. Biomarker concentrations were normalized (scale function in R) and the average scaled value is showed in blue and yellow scales. Blue shading represents the highest average scaled value; yellow shading, the lowest average scaled value. Each column (i.e., individual) in the heatmap is matched with color-coded cluster assignment: Cluster Control – green bar; Cluster Vivax^low^ – blue bar and Cluster Vivax^high^ – red bar.

In parallel to more severe thrombocytopenia in Vivax^high^ patients, plasma levels of cytokines inducing megakaryocytic differentiation in the bone marrow (BM), thrombopoietin (TPO) and IL-11, were significantly increased in this cluster (**Figure 4C, S4A**). In accordance with the pattern of immune cell fractions in the peripheral blood of *P. vivax* patients, the Vivax^high^ cluster showed a significant increase in the levels of pro-inflammatory cytokines associated with induction of myeloid-biased haematopoietic stem cell (HSC) differentiation and inhibition of lymphopoiesis in BM (e.g., TNF-α, IL-1α, IL-1β, IL-6, IL-8; **Figures 4C and S4B**) ^36–38^. In addition, Vivax^high^ patients had increased circulating levels of G-CSF, a major mediator of HSC-biased myelopoiesis and the neutrophil activation marker, L-Selectin (**Figures 4C and S4C**) ^40–42^. Together, these Luminex data support the haematological measurements, suggesting that a compensatory response is mounted in the BM to counterbalance the massive decrease of platelets in periphery. Upregulation of cytokines inducing myelopoiesis, while inhibiting lymphopoiesis ^36–38^, might also explain the decrease of lymphocyte counts and enrichment of activated neutrophils in the peripheral circulation of *P. vivax* patients.

### Elevated circulating markers of EC activation and damage in Vivax^high^ compared to Vivax^low^ patients

Patient clustering indicated that Vivax^high^ patients have increased levels of EC markers in the plasma compared to Vivax^low^ patients (**Figure 2C**). Previous studies indicate that EC activation and damage might contribute to thrombocytopenia and inducing hematopoiesis, resulting in HSC differentiation directed towards myelopoiesis ^23, 32, 33, 36–38, 43–45^. In our cohort, circulating levels of EC adhesion molecules (ICAM-1, VCAM-1, E-selectin and P-selectin) and other EC activation markers and procoagulant molecules (Ang-2, VWF-A2, CD40L and PAI-1) were significantly increased in the plasma of Vivax^high^ patients compared to Vivax^low^ patients and healthy controls (**Figures 5A, S5A, B**). Likewise, Syndecan-1, a marker of EC glycocalyx breakdown (i.e., damage of EC plasma membrane) ^46, 47^, was significantly increased Vivax^high^ but not in Vivax^low^ patients (**Figures 5A and S5C**).

**Figure 5:**
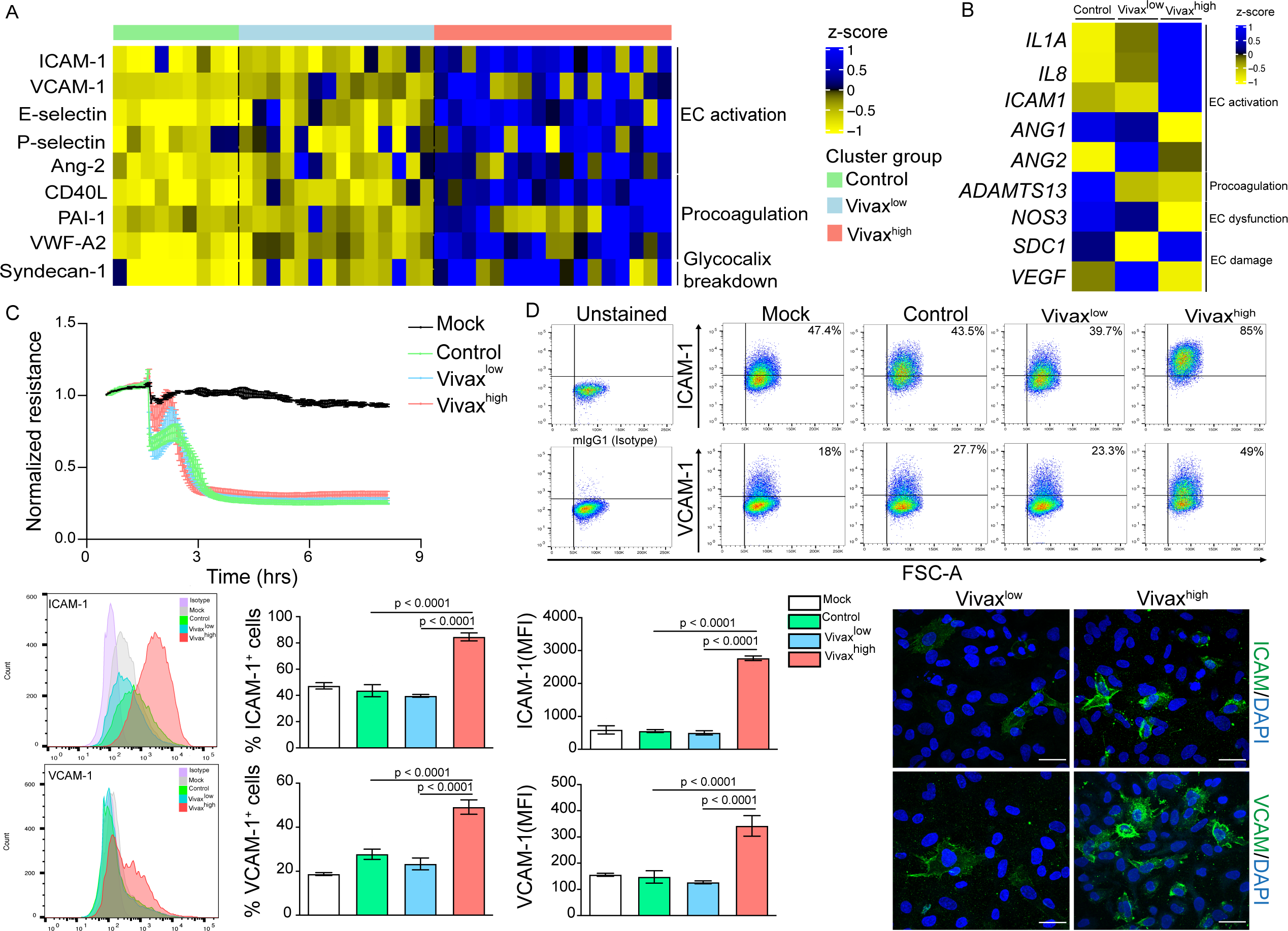
Elevated circulating markers of EC activation and damage in Vivax^high^ compared to Vivax^low^ patients. **(A) Endothelial changes across clusters: Luminex.** Heatmap represents z-scores obtained by centering values of Luminex data. Shown are markers of endothelial cell (EC) activation, procoagulation and glycocalyx damage. Each column (each individual) in the heatmap is matched with color-coded cluster assignment: Cluster Control – green bar; Cluster Vivax^low^ – blue bar and Cluster Vivax^high^ – red bar. **(B) Endothelial changes across clusters: qRT-PCR.** Transcriptional response of HUVECs incubated for 6h with pooled plasma from different clusters. Heatmap reflects relative mRNA expression intensity (average scaled value) after results were normalized to GAPDH housekeeping gene expression and untreated condition (mock). Data shown represent the mean of three independent experiments. For each experiment, 2 technical replicates were performed for each condition. **(C) Endothelial changes across clusters: impedance changes.** Endothelial monolayer integrity was measured during 20% v/v of pooled plasma incubation. Each line represents the mean ± SD of normalized resistance of HUVECs measured by ECIS at 4,000 Hz. Data shown are representative of three independent experiments. For each experiment, 2 technical replicates were performed for each condition. **(D) Endothelial changes across clusters: imaging and flow cytometry.** HUVECs were incubated for 18h with 30% v/v of pooled plasma of individuals in the different clusters or left untreated (mock). Percentage of cells expressing EC activation markers (adhesion molecules) ICAM and VCAM as well as quantification of protein expression was determined by flow cytometry and immunofluorescence analysis (scale bar = 33μM). Isotype antibodies were used as control to define positive populations. Significance was calculated for comparisons between conditions at the corresponding time point using. One-way analyses of variance statistical test with Tukey’s corrected multiple comparisons test. *p*-value is indicated above the graph when *p* < 0.05. Data shown are representative mean ± SEM of three independent experiments.

To independently test whether host factors and/or parasite products present in the plasma of the different patient groups can directly induce changes in ECs, we stimulated primary human umbilical vein endothelial cells (HUVECs) with pools of plasma from either Vivax^high^ patients, Vivax^low^ patients, or healthy controls. These experiments demonstrated that only pooled plasma from Vivax^high^ patients induces significant transcriptional upregulation of EC activation markers *ICAM-1*, *IL-1α*, *IL-8* along with downregulation of *Ang-1*, *ADAMTS13* and *NOS3* (eNOS) in HUVECs (**Figure 5B, S5D**). In contrast, expression of *Syndecan-1* and *VEGF*, two indicators of vascular damage, was not affected by either treatment (**Figure 5B, S5D**). Similarly, electric cell-substrate impedance sensing (ECIS) assays did not detect differences in functional perturbations in the endothelial cellular monolayer upon incubation with *P. vivax* pooled plasma when compared to control pooled plasma (**Figure 5C**). In contrast, flow cytometry and immunofluorescence assays performed with stimulated HUVECs revealed increased prevalence and protein expression levels of EC activation markers ICAM-1 and VCAM-1 upon exposure with Vivax^high^ pooled plasma **(Figure 5D, S5E)**, in support of qRT-PCR data. Together, these data demonstrate that *P*. *vivax* infection results in different ranges of EC alterations without massive cytoadhesion to these cells as observed in falciparum malaria. In addition, it indicates that local EC activation, mediated by direct or indirect interactions with parasitized RBCs, can be measured systemically.

### Indirect evidence for parasite-induced changes in deep tissues

To further investigate the interplay between host biomarkers and associated cellular responses as well as parasite parameters we constructed a network of interactions based on Pearson correlations with absolute correlation coefficient above 0.5 and *p*-value < 0.05 (**Figure 6A**). In addition, we also performed hierarchical clustering on matrices of Pearson correlations (*p*-value < 0.01) with selected modules of parasite and host parameters (**Figure 6B**). Data from Vivax^low^ and Vivax^high^ patient subgroups were combined for this analysis as they similarly contribute to the associations we found so far (**Figure S6**).

**Figure 6:**
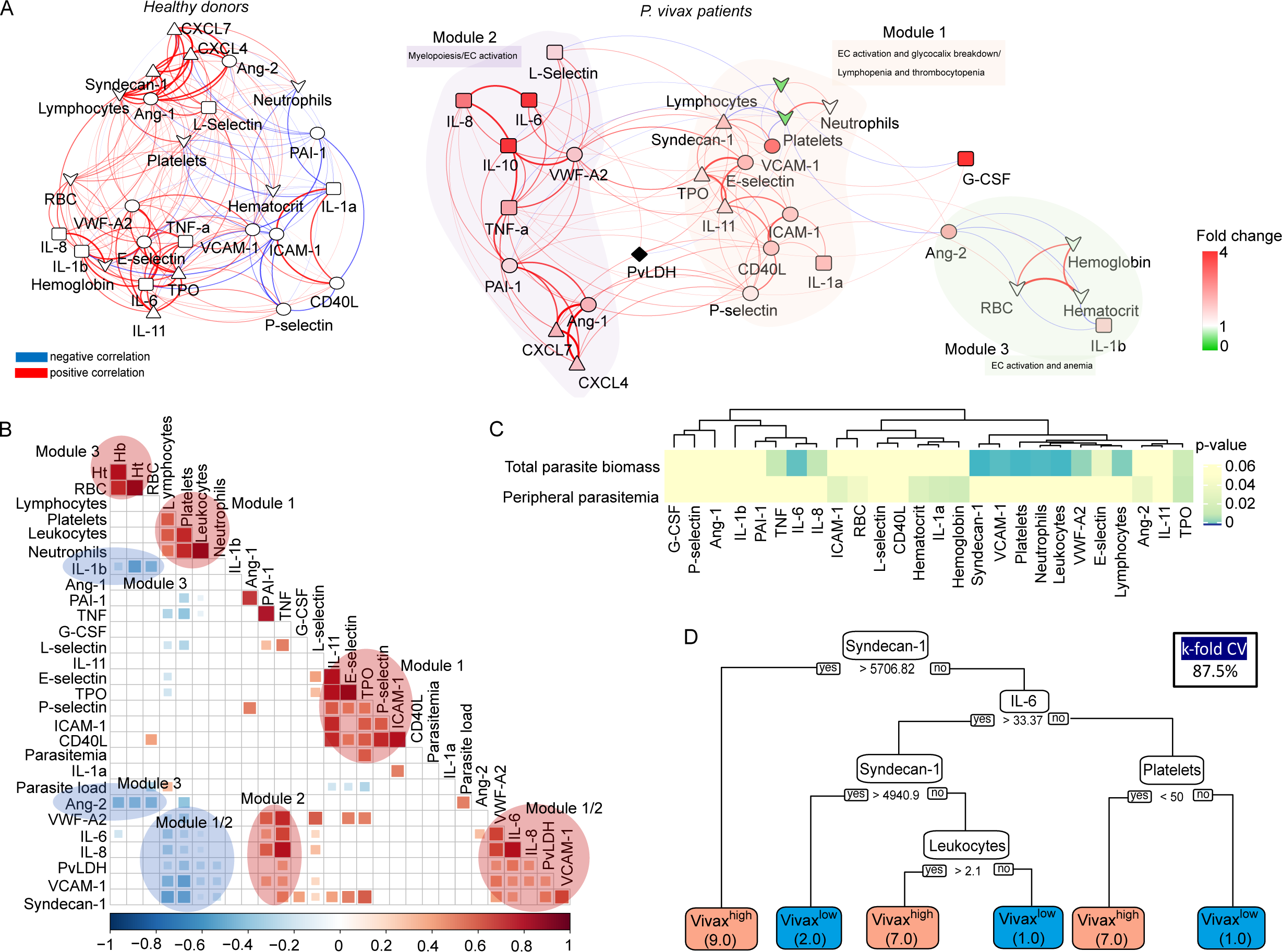
Network analysis and clustering of parasite and host signatures indicate parasite-induced changes in deep tissues. **(A) Network analysis.** Networks of the Pearson’s correlations (absolute coefficient above 0.5 and *p*-value < 0.05) between parasite biomass (PvLDH) and host signatures in healthy donors (left graph) and in *P. vivax*-infected patients (right graph), using a force-directed layout. The symbols of the nodes represent biological functions: triangle represents markers of platelet activation and thrombopoiesis-inducing cytokines; V shape represents haematological parameters (neutrophil, lymphocyte and platelet counts); circles represent endothelial cell activation markers; squares represent myelopoiesis-inducing cytokines and neutrophil activation markers. The colors in the nodes represent the fold change in relation to control levels. Because healthy donors do not have parasitemia, PvLDH node is represented in black. Each connecting line (edge) represents a significant interaction detected by the network analysis using R. Correlation strength is represented by edge color transparency and width. Positive correlations are represented by red edges and negatives correlations are represented by blue edges. **(B,C) Correlation matrix and heatmap. (B)** Representative image of Pearson’s correlation matrix calculated for all *P. vivax* patients. Only correlations with *p*-value <0.01 are shown and hierarchical clustering was applied. Red circles highlight positive correlations in the functional modules depicted in (A), and blue circles highlight negative correlations in the functional modules also depicted in (A). **(C)** Heatmap showing *p*-values of the correlations between different parasite parameters, parasite biomass (PvLDH) and peripheral parasitemia and host signatures (hematological and Luminex parameters). **(D) Decision tree model.** Best-fit classification tree model generated with the C4.5 algorithm showing Syndecan-1, IL-6 and platelet counts are the dominant variables capable to predict total parasite biomass in *P. vivax* patients. Cut-off values of the attribute that best divided groups were placed in the root of the tree according to the parameter value (pg/mL for soluble markers or number of cells x 1,000/μL of blood for platelet counts). The total of classified registers for each class are given in parentheses for each terminal node with the k-fold cross-validation (k-fold CV) accuracy indicated.

The network analysis revealed a decentralized topology, lower complexity and connectivity between the edges with data from *P. vivax* patients compared to the highly dense, homogenous and centralized network graph of healthy donors (91 edges vs 142 edges, respectively). Interestingly, the network graph of *P. vivax* patients is separated into three modules of strong interactions, with closely related biological functions. Module 1 is formed by markers of EC activation and damage, together with lymphocyte, platelet and neutrophil counts in addition to the megakaryocyte differentiation inducing cytokines (TPO and IL-11) (**Figure 6A**). In support of the role of EC activation and damage in the haematological changes observed in this cohort, hierarchical clustering revealed a positive correlation between adhesion molecules VCAM-1 and E-selectin and EC glycocalyx breakdown (Syndecan-1) (**Figure 6B**). In addition, VCAM-1, E-selectin, Ang-2 and VWF-A2, and Syndecan-1 are negatively correlated with platelet and lymphocyte counts, while ICAM-1 is positively correlated with neutrophil counts (**Figures 6A, B**). Module 2 is formed by proinflammatory cytokines with myelopoiesis inducing effects and molecules associated with platelet activation and coagulation cascades (**Figures 6A, B**). Interestingly, EC activation markers and Syndecan-1 (EC damage) from module 1 also display positive correlations with myelopoiesis-inducing cytokines from module 2 (**Figure 6B**). Finally, module 3 is formed by Ang-2 and the proinflammatory cytokine IL-1β associated with haemoglobin, haematocrit and RBC numbers (anemia markers) (**Figures 6A, B**). Most notably, PvLDH connects the two main functional modules 1 and 2 (**Figures 6A, B**). Accordingly with Figures 2 and 6A,B, the biological significance of total parasite biomass, but not peripheral parasitaemia or parasite load, in affecting host response is also corroborated by the high significant and positive associations of PvLDH with multiple host parameters, including Syndecan-1 (EC damage), VCAM-1, VWF (EC activation and platelet pooling), and IL-6, IL-8, TNF-α (inflammation and myelopoiesis inducing cytokines) (**Figure 6C**). Meanwhile, platelet, lymphocyte and neutrophil counts are negatively correlated with high significance (p-value < 0.0001) with total parasite biomass, but not with peripheral parasitaemia or parasite load (**Figure 6C**). Similar to Figure 2E, application of a best-fit classification tree model demonstrated that Syndecan-1, IL-6 and platelet counts are the most dominant predictor attributes capable to classify *P. vivax* patients based on total parasite biomass levels (**Figure 6D**). Using this model all *P. vivax* patients were correctly classified in either low (Vivax^low^) or high (Vivax^high^) total parasite biomass (PvLDH). In turn, PvLDH is a relevant predictor attribute (high information gain) in predicting thrombocytopenia severity and it is associated with increased severity of thrombocytopenia and lymphopenia in our cohort (**Figures S7 and S8**). Together, these data further support the hypothesis that a parasite population outside of circulation, as represented by total parasite biomass, is driving the host response including EC activation and damage as well as haematological disturbances (i.e., lymphopenia, thrombocytopenia and anaemia) in *P. vivax* patients **(Figures S7 and S8)**.

## Discussion

In this study, we performed a comprehensive analysis of host and parasite signatures detected in the plasma of a cross-sectional cohort of uncomplicated *P. vivax* malaria. Initial analysis of a series of circulating host biomarkers revealed significant levels of thrombocytopenia, lymphopenia and anaemia, as well as EC activation and damage across *P. vivax* patients compared to healthy controls. Deconvolution of heterogeneity across patients revealed two patient subgroups (Vivax^high^ and Vivax^low^) characterized by differences in total parasite biomass (based on circulating PvLDH levels) but not peripheral parasitaemia (based on blood smears). We observed a significant correlation between total parasite biomass (but peripheral parasitaemia) and systemic levels of markers of EC activation and damage and hematopoietic perturbations. In addition, by applying a supervised machine learning tree-structured model, we were able to associate EC damage and thrombocytopenia with parasite biomass. In agreement with a previous study ^8, 39^ our observations further suggest that total parasite biomass as measured by PvLDH is a better predictor of *P. vivax* host responses and pathogenesis than peripheral parasitaemia. Furthermore, these findings support the emerging paradigm of a major *P. vivax* parasite reservoir outside of circulation, in particular in the haematopoietic niche of bone marrow and spleen ^39^.

Thrombocytopenia, lymphopenia and anaemia are the most frequent *P. vivax* and *P. falciparum* associated haematological complications ^23–26^. In our cohort, 34%, 85% and 87% of patients exhibited anaemia, lymphopenia and thrombocytopenia, respectively. Although thrombocytopenia is not included in the World Health Organization (WHO) criteria for defining severe malaria, it has been associated with severe manifestations and the need for blood and platelet transfusions in severe vivax malaria, highlighting its clinical relevance in malaria diagnosis and treatment ^23, 24^. Various mechanisms have been proposed to explain the damage or excessive removal of platelets in *P. vivax* infection, including oxidative stress, platelet phagocytosis, IgG binding to platelet-bound malaria antigens, spleen pooling, or increased circulating nucleic acids levels ^23, 24, 48, 49^. EC activation and damage also play a role in intravascular platelet agglutination and increased platelet clearance from the circulation^28, 35^. Our data also demonstrate that thrombocytopenia is associated with an increase in IL-1, IL-6, IL-8, IL-10 and TNF-α. We also observed elevated levels of cytokines inducing megakaryocyte differentiation, TPO and IL-11, suggesting that a compensatory response is mounted in the BM to counterbalance the massive decrease of platelets in the periphery. In contrast, the relatively large drop in peripheral lymphocyte numbers we observed in the *P. vivax* patients is likely non-specific effect, e.g. pooling in the enlarged spleen rather than a response by *Plasmodium*-specific lymphocytes ^50^. Corroborating the potential role of total parasite biomass, rather than peripheral parasitaemia, in haematological disturbances (i.e., lymphopenia, thrombocytopenia and anaemia), Figures S7 and S8 show that total parasite biomass increases accordingly with thrombocytopenia and lymphopenia severity. Patients with severe thrombocytopenia also show more severe leukopenia, lymphopenia and mega platelets (higher MPV). In addition, plasma levels of cytokines, such as TNF-α, IL1-β, IL-8, IL-10; EC activation/damage markers, VCAM-1, E-selectin, VWF-A2, Ang-2, Ang-2:Ang1 ratio; Syndecan-1; thrombopoiesis-inducing cytokines, TPO and IL-11; platelet activation marker, CD40L and neutrophil activation marker, L-selectin, follow the increase in thrombocytopenia severity (**Figure S7**). A similar pattern is observed when stratifying patients based on lymphopenia severity (**Figure S8**). Interestingly, a tree-structured model demonstrated that PvLDH, along with VCAM-1 and Syndecan-1, is a relevant predictor attribute (high information gain) in predicting thrombocytopenia severity in our cohort (**Figure S7**).

Our data support previous studies suggesting a role for endothelial cell (EC) activation and damage in increased leukocyte adhesion, intravascular platelet agglutination with increased platelet clearance from the circulation and skewing of haematopoiesis toward the myeloid lineage (likely at the expense of lymphopoiesis) in the BM ^32–34, 36–38, 43, 44, 47^. *P. vivax* elicits a stronger inflammatory response and more pronounced endothelial activation when compared with other *Plasmodium* infections with similar or higher peripheral parasitaemia ^22^, however the role of EC activation in *P. vivax* pathogenesis is not yet understood. Damage of the EC plasma membrane, as represented by glycocalyx breakdown, has been associated with poor prognostic outcome in *P. falciparum* ^46^, but there is no data available for *P. vivax.* In our cohort soluble EC activation biomarkers (e.g., ICAM-1, VCAM-1, E-selectin, Ang-2, CD40L, vWF-A2) and the EC damage product, Syndecan-1, are positively correlated with thrombocytopenia, lymphopenia, anaemia and neutrophil enrichment in the peripheral blood. In addition, these biomarkers are positively correlated with increased circulating levels of cytokines inducing megakaryocyte-differentiation (e.g., IL-11 and TPO) and with cytokines inducing myeloid-biased HSC differentiation (e.g., TNF-α, IL1-α, IL6, IL-8 and G-CSF), suggesting both direct and indirect links between EC activation and damage and haematological perturbations. Total parasite biomass inducing EC activation might act synergistically with inflammatory changes potentially leading to splenic platelet pooling and platelet clumping in the vasculature without DIC ^20, 47, 51^. Likewise, increased activation-induced cell death (AICD) in T cells, splenic T-cell accumulation ^50^ or decreased lymphopoiesis due to myeloid-biased HSC differentiation induced by inflammatory cytokines and EC activation in the BM ^36, 37, 39^ might explain the severe lymphopenia and neutrophilia in vivax patients. Together such mechanisms could explain the link between parasite biomass and EC activation/damage with haematological changes observed in vivax patients that might contribute to pathogenesis and disease severity.

In a second series of experiments, we performed *ex vivo* stimulation of HUVECs with the plasma of the *P. vivax* cohort demonstrating that the mixture of parasite and host factors can directly induce EC activation in absence of parasitized RBCs. ECs are capable of responding to pathogens by sensing pathogen-associated molecular patterns (PAMPs) through pattern-recognition receptors (PRRs), which might play a key role in inducing EC activation when detecting *P. vivax* molecules enriched in the tissues where the parasite accumulates. ECs also express specific cytokine/chemokine receptors to detect proinflammatory signals released systemically or locally by activated immune cells in response to infection ^52, 53^. As result, EC activation induces exocytosis of secretory granules known as Weibel–Palade bodies that leads to the release of Ang-2 and VWF, as well as transcriptional programs that activate expression of adhesion molecules such as ICAM-1, VCAM-1, E-selectin, and secreted cytokines and chemokines ^33, 52, 53^. However, EC pathophysiology is complex, and changes represent a heterogenous spectrum ranging from simple perturbation to activation and even endothelial cell damage ^32^. Our Luminex data clearly confirm such heterogeneity in the spectrum of EC changes due to *P. vivax* infection, with systemic increase of markers of EC activation and damage only detected in Vivax^high^ patients. The *ex vivo* data show that increased systemic host proinflammatory factors and/or parasite products can alter EC properties, including activation of adhesion molecules and proinflammatory cytokines and downregulation of ADAMTS13. In contrast, vascular integrity was not affected. These data indicate that systemic inflammatory signatures in *P. vivax* patients can lead to local EC activation but not vascular damage, central events in malaria pathogenesis.

Although our study lacks longitudinal information, the findings might have clinical implications during and after treatment of vivax malaria. Several case reports demonstrate progressive clinical deterioration after commencement of treatment in *P. vivax* patients, associated with parasite killing that results in haemolysis of iRBCs and intravascular inflammation and oedema in response to the products released from these cells ^54–57^. Patients presenting with a strong host response during acute infection might therefore be at increased risk of deteriorating and developing severe symptoms after commencement of treatment (**Figure S9**). Thus, identification of unique biological signatures in *P. vivax* patients might help to build rational approaches to the diagnosis, prognosis and individualized treatment to modulate the host response to vivax malaria.

## Conclusions

Altogether, our data indicate that changes in clinical parameters and biomarkers detected in the plasma of *P. vivax* patients are the result of both, systemic host responses and local infection in tissue reservoirs such as BM and spleen. Our analysis shows that measuring a combination of host parameters (e.g., Syndecan-1, IL-6, platelet levels) and total parasite biomass (PvLDH) could predict the extent of parasite infection outside of circulation. Our data also instigate future investigations of systemic signatures with parallel analysis focused on tissue responses, in particular in reservoirs such as the haematopoietic niche of BM and spleen, which has great potential to advance better diagnosis and treatment of *P. vivax*.

### Ethics Approval and Consent to Participate

All subjects enrolled in the study were adults. Written informed consent was obtained from all participants and the study was conducted according to the Declaration of Helsinki principles. The study was approved by the local Research Ethics Committee at Fundação de Medicina Tropical Dr. Heitor Vieira Dourado (FMT-HVD, Manaus, Brazil), under #CAAE: 54234216.1.0000.0005.

## Data Availability

The data that support the findings of this study are available from the corresponding author upon reasonable request.

## Acknowledgements

We would like to thank all patients enrolled in this research and the support of the malaria diagnosis and field team at field team in the Fundação de Medicina Tropical Dr. Heitor Vieira Dourado (FMT-HVD) in Manaus, Brazil. The authors also gratefully acknowledge the help and assistance provided by the Central Laboratory of High-Performance Technologies (LaCTAD, University of Campinas) and the Institute of Infection, Immunity and Inflammation Flow Core Facility in the generation of some of the data reported in this manuscript. M.M. is supported by a Wolfson Merit Award from the Royal Society and Wellcome Trust Center award (number 104111). J.LS.F. was supported by the Sao Paulo Research Foundation (FAPESP grant 2019/01578-2 and 2016/12855-9), and F.T.M.C is supported by the Sao Paulo Research Foundation (FAPESP grant 2017/18611-7). M.V.G.L. and F.T.M.C. are CNPq research fellows.

## Conflict of interest

The authors have declared that no conflict of interest exists.

## Author Contributions

Designing research studies: JLSF, JCKS, CJ, EVP, SCPL, MM, FTMC.

Conducting experiments: JLSF, JCKS, CJ, KV.

Acquiring data: JLSF, JCKS, CJ, KV, BD, DL, HN.

Analysing data: JLSF, JCKS, DB, DL, HN.

Providing reagents: EVP, MVGL, MM, FTMC

Project administration: MM, FTMC

Writing – original draft: JLSF, MM, FTMC.

Writing – review & editing: JLSF, JCKS, DB, SCPL, MVGL, MM, FTMC

## Supporting Information

**Figure S1: Demographic and clinical features of all *P. vivax*-infected patients compared with selected 31 patients for multiplex-bead based assay and downstream analysis.** (A) Gender, age and hematological parameters compared between all 79 *P. vivax*-infected patients (All) and those 31 selected for downstream molecular analysis. **(B)** Comparison of gender, age and hematological parameters between 31 selected (S) *P. vivax*-infected patients with the remaining 48 non-selected (NS) patients. Parameters are depicted as box plots showing each individual value and the median with maximum and minimum values.

**Figure S2: Principal components analysis metrics. (A, B)** Analysis of eigenvalues (measure of the amount of variation retained by each principal component) and the percentage of explained variances retained by the PCs demonstrated that the first 10 PCs accounted for the variance of the data. **(C)** However, most of the variables were highly represented in the first 2 PCs (Dim 1 and Dim 2), which were therefore retained for further analysis.

**Figure S3: Methods determining the number of clusters best representing the data. (A, B)** PCA plots indicating different K-means cluster configurations, using k= 3 and k=4 clusters, respectively after performing bootstrapping. With k=3 different starting points give different clusters. The two most common clusters (top row) are very similar and they are obtained in 241 and 179 starts out of 1,000, respectively. However, the clustering that best represents the data when k = 3 is the third one found in 168/1000 starting points as its withinss (wss) metric is lower (highlighted in red). Indeed, this configuration is more equivalent to those clustering configurations when k=2. **(B)** Clusters seem more stable when k=4. Accordingly, the best clustering appears to be the ones represented in the bottom row, which contains two main groups and two small groups with just 2 patients. **(C, D)** The second method used was the Monte Carlo Reference-based Consensus Clustering (M3C), which also indicated that k=2 is the optimal number of clusters, as indicated in **(C)** the flat line in the CDF plot and **(D)** in the highest Relative Cluster Stability Index (RCSI) plot. **(E, F, G).** Using spectral clusters, instead of elliptical K-means clusters, M3C analysis indicates that k=3 gives the optimal number of clusters, as indicated in the **(E)** CDF plot, **(F)** RCSI plot and **(G)** the NXN consensus matrix, where each element represents the fraction of times two samples clustered together.

**Figure S4: Increase of thrombopoiesis- and myelopoiesis inducing cytokines in the plasma of Vivax^high^ patients. (A)** Levels of myelopoiesis-inducing cytokines, **(B)** thrombopoiesis-inducing cytokines thrombopoietin (TPO) and IL-11 and **(C)** neutrophil activation markers in the acute-phase patients’ plasma samples of our cross-sectional cohort in Manaus, Brazil, were determined by multiplex-bead based assay (Luminex): Control (healthy donors, n = 9), Vivax^low^ patients (n=14) and Vivax^high^ patients (n=17). Biomarkers’ concentration is depicted as scatter plots showing individual data points and the median (black lines) and the interquartile range. One-way analyses of variance with Bonferroni-corrected multiple comparisons test were performed. p-value is indicated above the graph when reached p < 0.05.

**Figure S5: Increase of markers of endothelial cell activation, damage (glycocalyx breakdown) and procoagulation in the plasma of Vivax^high^ patients. (A)** Levels of EC activation markers, **(B)** procoagulant phenotype and **(C)** EC damage (glycocalyx breakdown) in the acute-phase patients’ plasma samples of our cross-sectional cohort in Manaus, Brazil, were determined by multiplex-bead based assay (Luminex): Control (healthy donors, n = 9), Vivax^low^ patients (n=14) and Vivax^high^ patients (n=17), as indicated in the legend (top right corner). Biomarkers’ concentration is depicted as scatter plots showing individual data points and the median (black lines) and the interquartile range. One-way analyses of variance with Bonferroni-corrected multiple comparisons test were performed. p-value is indicated above the graph when reached significance of p < 0.05. **(D)** Quantitative mRNA expression was determined by qRT-PCR in RNA extracted from HUVECs incubated for 6h with 30% v/v pooled-plasma of individuals in the different clusters, as indicated in the legend (top right corner). Graphs depict relative expression after results were normalized to GAPDH housekeeping gene expression. The data shown are mean ± SEM representative of three independent experiments. Significance was calculated for comparisons between conditions at the corresponding time point using One-way analyses of variance with Tukey’s corrected multiple comparisons test. p-value is indicated above the graph when reached significance of p < 0.05. **(E)** Schematics of HUVECs gating strategy used for flow cytometry analysis. Endothelial cells gate (ECs) was defined based on the cells’ forward scatter (FSC) and side scatter (SSC). Further, gated single cells on FSC-A vs FSC-H scatter plot and selected live cells based on Fixable Viability Dye eFluor™ 506 staining.

**Figure S6: Representative images of Pearson’s correlation matrix calculated separately for each *P. vivax* patient cluster. (A)** Vivax^low^ patients (n=14). **(B)** Vivax^high^ patients (n=17). A reduced complexity model was established by focusing on informative interactions between *P. vivax* and host signatures determined by Pearson’s correlation coefficients. Only correlations with associated *p*-value <0.01 are shown and hierarchical clustering was applied.

**Figure S7: Validation of patients’ clusters and correlations when segregating patients based on thrombocytopenia severity.** Box plots showing **(A)** parasite parameters, clinical parameters and biomarkers measured on plasma samples were generated segregating patients based on levels of thrombocytopenia severity (normal, mild, moderate and severe) color-coded. **(B)** Recursive partitioning classification tree model generated with the rpart function in R showing the high value of VCAM-1, PvLDH and Syndecan-1 to predict thrombocytopenia severity in *P. vivax* patients. Cut-off values of the attribute that best divided groups were placed in the root of the tree according to the parameter value (pg/mL for soluble markers or O.D. for PvLDH).

**Figure S8: Validation of patients’ clusters and correlations when segregating patients based on lymphopenia severity.** Box plots showing parasite parameters, clinical parameters and biomarkers measured on plasma samples were generated segregating patients based on levels of lymphopenia severity (normal, moderate and severe) color-coded.

**Figure S9: Haemolysis potentiates Vivax^high^-induced EC activation.** In order to mimic the environment associated with commencement of anti-malarial treatment, such as content released from haemolysis of RBCs and dead parasites biproducts, HUVECS were stimulated with either *P. vivax*-infected (schizont enriched) or uninfected RBCs lysates in 30% v/v pooled-plasma of individuals in the different clusters. For the parasite lysates, batch pellets of *P. vivax* iRBCs enriched of schizonts, isolated from blood of P. vivax-infected patients using Percoll gradient centrifugation, and healthy donor RBCs were stored at −80 °C without any cryopreservative agent. Pellets were twice freeze–thawed for use as whole lysates. Total RNA was extracted 6h after treatment and relative mRNA expression determined by real-time quantitative PCR. Graphs depict relative expression after results were normalized to GAPDH housekeeping gene expression. The data shown are mean ± SEM representative of three independent experiments. Significance was calculated for comparisons between conditions at the corresponding time point using One-way analyses of variance with Tukey’s corrected multiple comparisons test. p-value is indicated above the graph when reached significance of p < 0.05. Haemolysis of either *P-vivax*-infected or uninfected RBCs potentiates the effect of Vivax^high^ pooled plasma in inducing transcriptional upregulation of EC activation markers. Different from the stimulation only with plasma, in the presence of haemolysis, we also observed upregulation of *Ang-2* and *VEGF*, and downregulation of *NOS3*, markers associated with perturbation of the vascular integrity and function.

**Table S1:** Measurements of K-means cluster stability, using k=2, k=3 and k=4 clusters, via bootstrapping. The metrics of interest is jaccard_index which measures the cluster similarity across bootstrap samples. Jaccard_index ranges from 0 to 1, an index < 0.6 hints at a weak, unreliable cluster while > 0.85 means generally reliable.

| kmeans_bootstrap |  |  |  |  |  |
| --- | --- | --- | --- | --- | --- |
| n_clusters | seed | withinss | jaccard_index | cluster_id | cluster_size |
| 2 | 1 | 1122.0 | 0.903 | 1 | 21 |
| 2 | 1 | 1122.0 | 0.860 | 2 | 18 |
| 3 | 9 | 994.6 | 0.860 | 1 | 21 |
| 3 | 9 | 994.6 | 0.726 | 2 | 14 |
| 3 | 9 | 994.6 | 0.539 | 3 | 4 |
| 3 | 3 | 996.6 | 0.852 | 1 | 19 |
| 3 | 3 | 996.6 | 0.702 | 2 | 16 |
| 3 | 3 | 996.6 | 0.559 | 3 | 4 |
| 3 | 14 | 994.4 | 0.861 | 1 | 21 |
| 3 | 14 | 994.4 | 0.757 | 2 | 16 |
| 3 | 14 | 994.4 | 0.442 | 3 | 2 |
| 3 | 1 | 1028.2 | 0.693 | 1 | 16 |
| 3 | 1 | 1028.2 | 0.297 | 2 | 5 |
| 3 | 1 | 1028.2 | 0.834 | 3 | 18 |
| 3 | 4 | 1031.1 | 0.496 | 1 | 14 |
| 3 | 4 | 1031.1 | 0.499 | 2 | 9 |
| 3 | 4 | 1031.1 | 0.680 | 3 | 16 |
| 3 | 28 | 997.7 | 0.425 | 1 | 2 |
| 3 | 28 | 997.7 | 0.852 | 2 | 19 |
| 3 | 28 | 997.7 | 0.712 | 3 | 18 |
| 4 | 4 | 902.0 | 0.679 | 1 | 12 |
| 4 | 4 | 902.0 | 0.806 | 2 | 18 |
| 4 | 4 | 902.0 | 0.612 | 3 | 4 |
| 4 | 4 | 902.0 | 0.427 | 4 | 5 |
| 4 | 9 | 907.3 | 0.529 | 1 | 12 |
| 4 | 9 | 907.3 | 0.588 | 2 | 9 |
| 4 | 9 | 907.3 | 0.609 | 3 | 4 |
| 4 | 9 | 907.3 | 0.701 | 4 | 14 |
| 4 | 10 | 893.2 | 0.579 | 1 | 2 |
| 4 | 10 | 893.2 | 0.459 | 2 | 7 |
| 4 | 10 | 893.2 | 0.631 | 3 | 11 |
| 4 | 10 | 893.2 | 0.795 | 4 | 19 |
| 4 | 74 | 905.9 | 0.664 | 1 | 15 |
| 4 | 74 | 905.9 | 0.509 | 2 | 13 |
| 4 | 74 | 905.9 | 0.610 | 3 | 9 |
| 4 | 74 | 905.9 | 0.608 | 4 | 2 |
| 4 | 12 | 885.9 | 0.813 | 1 | 19 |
| 4 | 12 | 885.9 | 0.470 | 2 | 2 |
| 4 | 12 | 885.9 | 0.596 | 3 | 2 |
| 4 | 12 | 885.9 | 0.650 | 4 | 16 |
| 4 | 20 | 883.9 | 0.760 | 1 | 21 |
| 4 | 20 | 883.9 | 0.458 | 2 | 2 |
| 4 | 20 | 883.9 | 0.715 | 3 | 14 |
| 4 | 20 | 883.9 | 0.610 | 4 | 2 |

**Table S2:** Correlation (loading score) of variables to principal components.

| **PC1** | **correlation** | **p.value** |
| --- | --- | --- |
| VWF.A2 | 0.8256501 | 9.90E-11 |
| TPO | 0.8191482 | 1.84E-10 |
| TNF | 0.8141927 | 2.90E-10 |
| IL.11 | 0.7827225 | 3.93E-09 |
| E.selectin | 0.7822878 | 4.06E-09 |
| CD40L | 0.7270363 | 1.59E-07 |
| Syndecan.1 | 0.7224769 | 2.07E-07 |
| L.Selectin | 0.6730125 | 2.68E-06 |
| PAI.1 | 0.5994172 | 5.52E-05 |
| pvLDH | 0.5850459 | 9.17E-05 |
| P.selectin | 0.5774233 | 1.19E-04 |
| Parasitemia | 0.5650752 | 1.78E-04 |
| IL.8 | 0.5544242 | 2.50E-04 |
| VCAM.1 | 0.5410419 | 3.76E-04 |
| IL.10 | 0.5352827 | 4.46E-04 |
| ICAM.1 | 0.5188345 | 7.14E-04 |
| Ang.2 | 0.489315 | 1.57E-03 |
| IL.6 | 0.4463894 | 4.39E-03 |
| NF | 0.3841315 | 1.58E-02 |
| IL.1a | 0.3820866 | 1.64E-02 |
| IL.1b | 0.3649609 | 2.23E-02 |
| CXCL7 | 0.324584 | 4.38E-02 |
| Lymp | -0.7239061 | 1.91E-07 |
| Plt | -0.7614057 | 1.82E-08 |
| **PC2** | **correlation** | **p.value** |
| Ang.1 | 0.8788255 | 1.90E-13 |
| CXCL4 | 0.8736698 | 3.92E-13 |
| CXCL7 | 0.8667025 | 9.94E-13 |
| PAI.1 | 0.6289549 | 1.80E-05 |
| P.selectin | 0.3833705 | 1.60E-02 |
| IL.8 | 0.3543761 | 2.69E-02 |
| TNF | 0.3541656 | 2.70E-02 |
| MPV | -0.3162224 | 4.99E-02 |
| Syndecan.1 | -0.3166453 | 4.95E-02 |
| IL.1a | -0.3530831 | 2.75E-02 |
| **PC3** | **correlation** | **p.value** |
| Ht | 0.8587232 | 2.72E-12 |
| RBC | 0.7727747 | 8.21E-09 |
| Hb | 0.7288971 | 1.43E-07 |
| ICAM.1 | 0.5463348 | 3.21E-04 |
| CD40L | 0.4927518 | 1.44E-03 |
| IL.11 | 0.4018958 | 1.12E-02 |
| MPV | 0.3724087 | 1.96E-02 |
| IL.8 | -0.3186183 | 4.81E-02 |
| Ang.2 | -0.3549913 | 2.66E-02 |
| IL.6 | -0.3628678 | 2.32E-02 |
| SCF | -0.4315038 | 6.09E-03 |

**Table S3:**
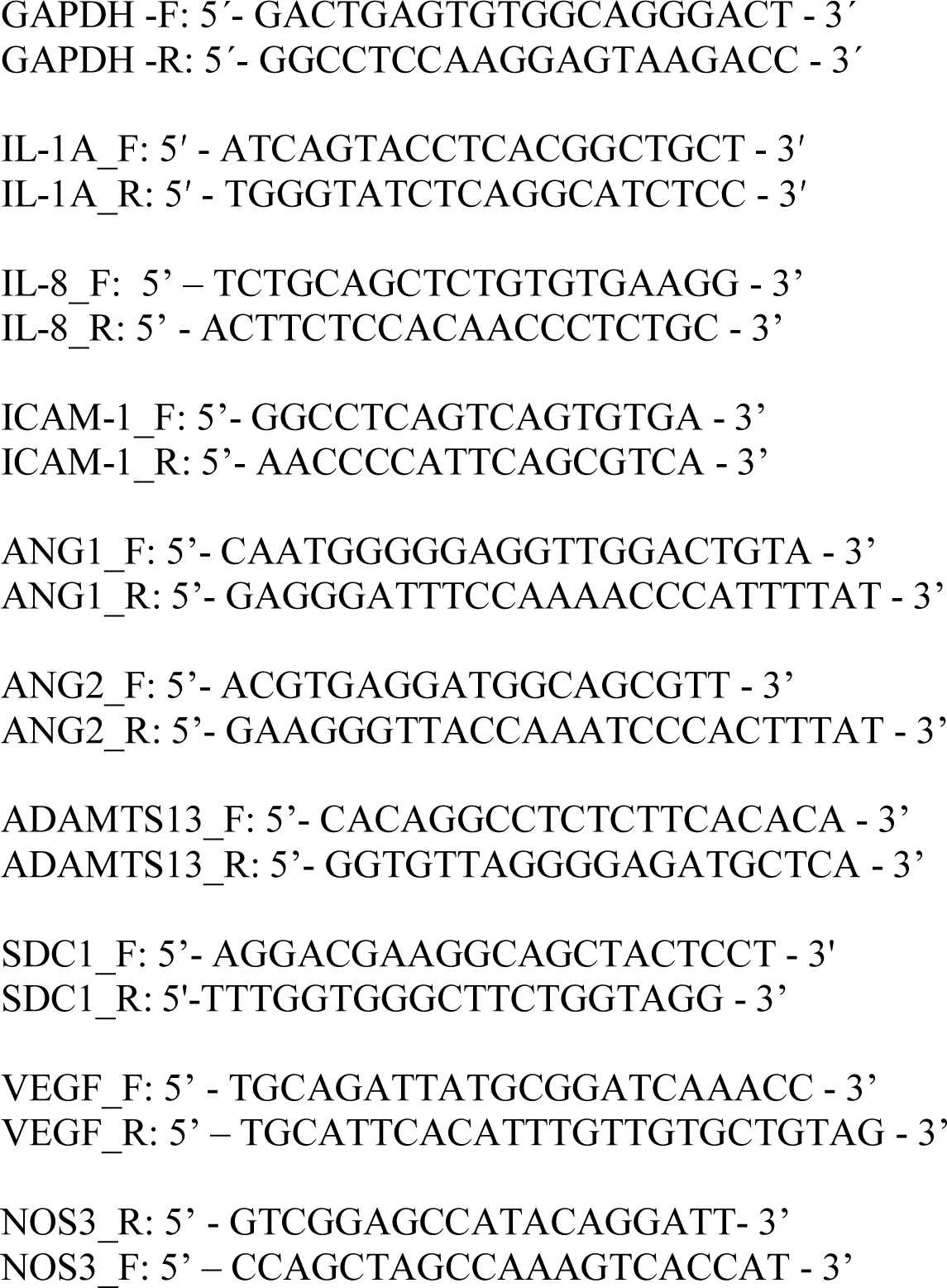
Oligo sequences used in the qRT-PCRs.

